# Type 2 Diabetes Partitioned Polygenic Scores Are Differentially Associated with Aging Hallmarks

**DOI:** 10.64898/2026.07.06.26357294

**Authors:** Masashi Hasebe, Chen-Yang Su, Chi Zhao, Tianyuan Lu, Cassandra N. Spracklen, Satoshi Yoshiji

## Abstract

Type 2 diabetes (T2D) arises from distinct diabetogenic mechanisms, but whether these mechanisms differ in their associations with hallmarks of aging remains unclear. We analyzed 449,505 UK Biobank and 374,973 *All of Us* participants using an overall T2D polygenic score (oPS) and eight partitioned polygenic scores (pPSs) representing distinct T2D-related mechanisms. Across organ systems, 81 age-related diseases were assigned to nine hallmarks of aging. UK Biobank analyses used Cox regression for incident hallmark-level outcomes, and *All of Us* analyses used logistic regression for prevalent hallmark-level outcomes. In both cohorts, the oPS was associated with disease burden across hallmarks, whereas pPS associations varied by mechanism. The obesity pPS showed the strongest and most consistent associations, while other insulin-resistance-related pPSs, including the lipodystrophy pPS, showed more modest positive associations. Beta-cell dysfunction pPS associations were close to null across hallmarks. Obesity pPS-hallmark associations were significantly attenuated after adjustment for BMI, and lipodystrophy pPS-hallmark associations after adjustment for triglyceride-to-HDL cholesterol ratio (TG/HDL-C), a marker of insulin resistance. These findings suggest that adiposity and insulin resistance, indexed by BMI and TG/HDL-C, may act as modifiable factors in the T2D genetic burden on aging hallmarks.

**Graphical Abstract:** 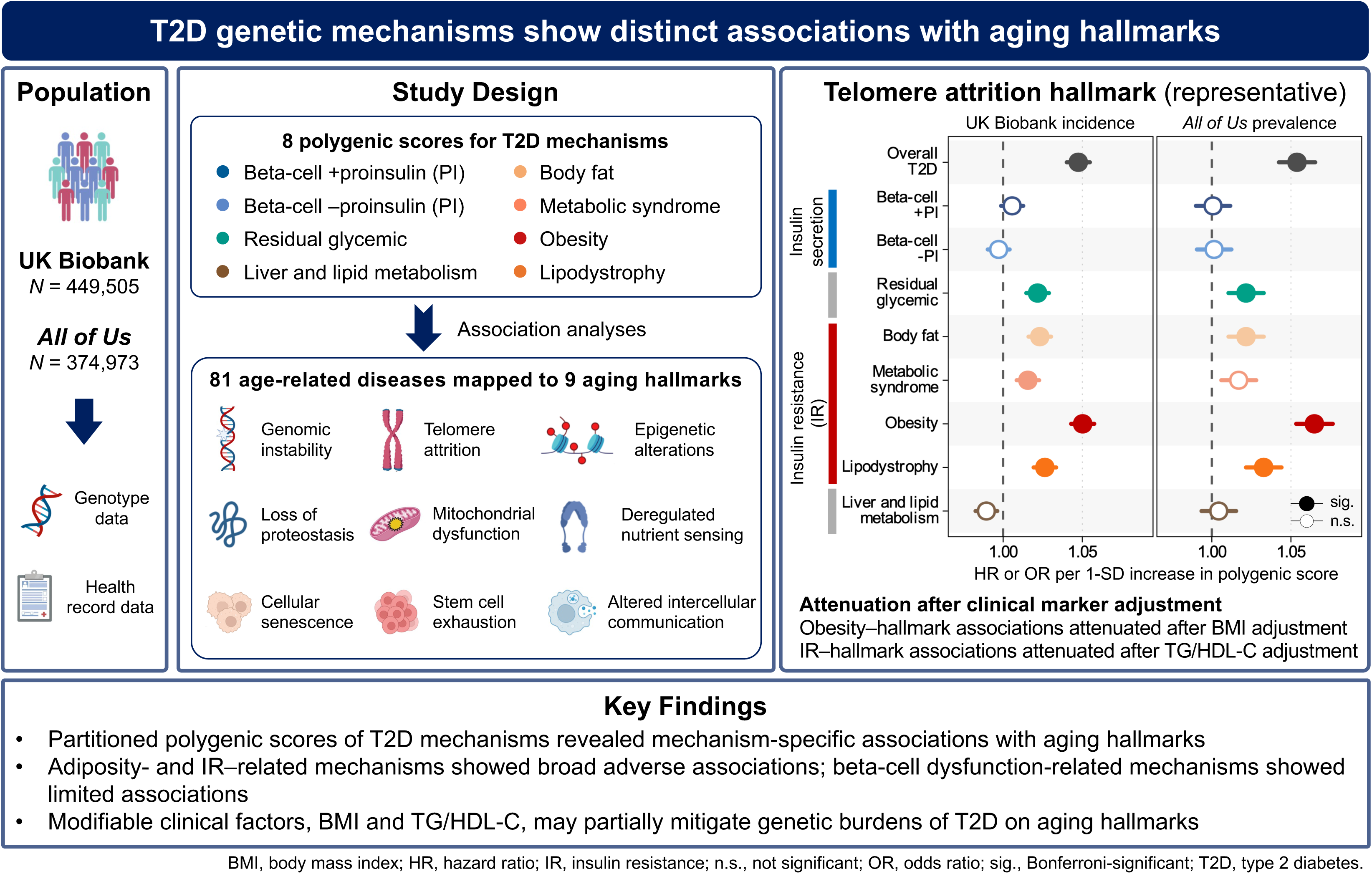

## INTRODUCTION

Type 2 diabetes (T2D) is biologically and clinically heterogeneous, reflecting distinct diabetogenic mechanisms such as beta-cell dysfunction, excess adiposity, and insulin resistance (1–4). People with T2D also experience accelerated aging and an increased burden of age-related morbidity, including cardiovascular disease, neurodegenerative disease, and cancer (5, 6). However, whether distinct mechanisms of T2D are differentially related to aging and age-related morbidity remains unclear.

Recent genetic studies have helped clarify the mechanistic heterogeneity of T2D (7). In 2024, the largest multi-ancestry genome-wide association study (GWAS) of T2D grouped 1,289 associated variants into biologically informed clusters representing distinct underlying processes, including mechanisms related to beta-cell dysfunction, adiposity, and insulin resistance (2). Cluster-specific partitioned polygenic scores (pPSs) derived from these clusters have been associated with differences in clinical presentation, disease progression, and diabetes-related complications (2, 8–10). More recently, these pPSs have also been linked to selected outcomes beyond conventional diabetes-related complications (11, 12). However, their associations with a broader spectrum of age-related diseases have not been systematically evaluated.

Age-related diseases are heterogeneous, frequently co-occur, and span multiple organ systems, so analyses of individual diseases may miss broader patterns of age-related morbidity (13, 14). The hallmarks of aging framework addresses this complexity by organizing biological processes implicated in aging into nine domains: genomic instability, telomere attrition, epigenetic alterations, loss of proteostasis, deregulated nutrient sensing, mitochondrial dysfunction, cellular senescence, stem cell exhaustion, and altered intercellular communication (15, 16). Using this framework, previous studies have mapped clinically manifest age-related diseases in electronic health record (EHR) data to hallmark-related categories, enabling biologically informed grouping of outcomes in population research (14, 17). Large-scale studies have further shown that exposures such as obesity and social disadvantage are associated with distinct patterns of hallmark-related disease risk (18, 19). Given that several mechanisms central to T2D have been implicated in aging (20–22), pPSs may also show distinct associations with clinically manifest age-related diseases grouped according to the hallmarks of aging. Examining these associations may clarify whether age-related disease burden linked to T2D differs across diabetogenic mechanisms and whether these differences relate to modifiable cardiometabolic traits.

In this study, we examined the associations of cluster-specific T2D pPSs, together with an overall polygenic score (oPS) for T2D representing aggregate genetic liability, with hallmark-specific age-related diseases in the UK Biobank and then assessed the consistency of these associations in the *All of Us* Research Program. We aimed to determine whether genetically defined components of T2D heterogeneity show distinct and consistent associations across these hallmark-defined disease groups, with particular attention to mechanisms related to adiposity and insulin resistance.

## RESEARCH DESIGN AND METHODS

### Data Sources

Analyses were conducted in the UK Biobank, a prospective cohort of >500,000 participants recruited at ages 40–69 years from 22 U.K. assessment centers between 2006 and 2010 (23). We included participants genotyped on the UK BiLEVE or UK Biobank Axiom arrays with centrally quality-controlled imputed genotype data based on the Haplotype Reference Consortium, UK10K, and 1000 Genomes reference panels (24). We excluded participants who withdrew consent or had sex discordance, sex chromosome aneuploidy, excess heterozygosity or missingness, missing genotyping batch information, or second-degree or closer relatedness (kinship score ≥0.0884).

For external comparison, we conducted analogous analyses in the *All of Us* Research Program, a large and diverse U.S. cohort launched by the National Institutes of Health in 2018 (25). We used data from the Curated Data Repository version 8 for the Controlled Tier, including participants with linked EHR data and available short-read whole-genome sequencing (srWGS) data generated from blood-derived DNA on the Illumina NovaSeq 6000 platform under standardized procedures (26). We excluded participants with low-quality srWGS data and those with close relatedness as defined in the relatedness table provided with the release (kinship score >0.1).

The UK Biobank received ethical approval from the North West Multi-centre Research Ethics Committee (REC reference 11/NW/0382), and the *All of Us* Research Program was approved by the *All of Us* Institutional Review Board. All participants in both cohorts provided written informed consent at recruitment.

### Polygenic Score Calculation

We constructed an oPS and eight pPSs using allelic weights derived from the largest multi-ancestry GWAS meta-analysis of T2D (2). Because the original GWAS included participants of European and South Asian ancestry from the UK Biobank, we used weights derived from a UK Biobank-excluded version of the meta-analysis to minimize potential bias from sample overlap. The oPS used all 1,289 independent genome-wide significant index variants from the original meta-analysis, whereas each pPS used the subset assigned to one of the reported mechanistic clusters (2): two beta-cell dysfunction clusters primarily reflecting impaired insulin secretion, with and without proinsulin (beta cell +PI and beta cell −PI, respectively); four insulin-resistance-related clusters (body fat, metabolic syndrome, obesity, and lipodystrophy); and two clusters without clear predominance of either process (residual glycemic and liver and lipid metabolism) (**Supplementary Table 1**). Scores were calculated in PLINK v2.0 as weighted sums of T2D risk-allele dosages using imputed dosages in the UK Biobank and srWGS data in *All of Us* (27). In each cohort, scores were based on available variants. For ancestry-stratified analyses, ancestry-specific PSs were calculated using corresponding ancestry-specific GWAS weights; for European and South Asian strata, these weights were derived from summary statistics excluding UK Biobank participants.

### Ascertainment of Hallmark-Specific Age-Related Diseases and Mortality

In the UK Biobank, hallmark-specific age-related diseases were ascertained from nationally linked inpatient hospital and death registry records using ICD-10-coded diagnoses and corresponding event dates. ICD-10 codes were grouped into the nine hallmarks of aging using definitions based on the hallmark-disease taxonomy of Kuan et al. (17) and Fraser et al. (14), together with the ICD-10 implementation used in prior multicohort studies (18, 19). In the original framework, each hallmark comprises 27–30 diseases, with some diseases assigned to more than one hallmark, so hallmark-level disease groups are not mutually exclusive. Because T2D genetic liability was the exposure of interest, diabetes-related ICD-10 codes (E10-E14) were excluded, leaving 81 diseases (**Supplementary Table 2**).

In *All of Us*, hallmark-specific age-related diseases were ascertained among participants who consented to EHR data sharing and had available EHR data in the Observational Medical Outcomes Partnership (OMOP) Common Data Model (28). Outcome ascertainment was restricted to inpatient diagnoses. To operationalize hallmark-disease definitions originally curated in ICD-10, we mapped them to standard OMOP condition concepts and compiled the resulting implementation table (**Supplementary Table 3**). Matching inpatient diagnoses were then classified as hallmark-specific age-related diseases. Additional details are provided in **Supplementary Note 1**.

### Statistical Analysis

Primary analyses were performed separately in each cohort using all eligible participants. To facilitate comparison across scores and cohorts in the primary analyses, the eight pPSs and the oPS were residualized on the top 10 genetic principal components (PCs) and standardized within each cohort. Ancestry-stratified analyses were conducted as sensitivity analyses.

We first examined associations of the PSs with cardiometabolic traits to assess their consistency with the cluster-specific cardiometabolic patterns reported in the source GWAS (2). Associations were evaluated using linear regression adjusted for age, sex, and the top 10 genetic PCs in both cohorts, with additional adjustment for assessment center and genotyping array in the UK Biobank. Detailed trait definitions and analytic procedures are provided in **Supplementary Note 2**.

We then examined associations of the PSs with hallmark-defined groups of age-related diseases. Analyses focused on incident outcomes in the UK Biobank and prevalent outcomes in *All of Us*, where harmonized longitudinal follow-up for inpatient outcomes was shorter. In the UK Biobank, incident outcomes were defined as the first post-baseline occurrence of any disease assigned to a given hallmark, including death attributable to a disease within the same hallmark. In *All of Us*, prevalent outcomes were defined as at least one disease assigned to a given hallmark at or before baseline. Within-hallmark multimorbidity was defined as the occurrence of a second or third distinct disease within the same hallmark in UK Biobank incident analyses, and as at least two or at least three distinct diseases within the same hallmark in *All of Us* prevalent analyses.

Incident outcomes in the UK Biobank were analyzed using Cox proportional hazards models after excluding participants who had already met the corresponding outcome definition at baseline. Follow-up extended from baseline until the outcome of interest, death, loss to follow-up, or administrative censoring at the end of follow-up in England (31 March 2023), Scotland (31 August 2022), or Wales (31 May 2022), whichever came first. The proportional hazards assumption was assessed using scaled Schoenfeld residuals. Fine-Gray subdistribution hazard models were additionally used, treating death not attributable to diseases within the corresponding hallmark as a competing event (29). Prevalent outcomes in *All of Us* were analyzed using logistic regression. To further characterize disease-specific patterns underlying hallmark-level associations, individual age-related diseases were evaluated separately in UK Biobank incident analyses. All models were adjusted for age at baseline, sex, and the top 10 genetic PCs in both cohorts, with additional adjustment for assessment center and genotyping array in the UK Biobank.

Because PS distributions and disease associations may still vary across ancestry groups after PC adjustment (30), ancestry-stratified sensitivity analyses were performed using ancestry-specific PSs. These analyses focused on the cohort-specific hallmark-level outcomes defined above. Ancestry-stratified estimates were pooled within each cohort using DerSimonian-Laird random-effects meta-analysis (see **Supplementary Note 3**) (31).

To assess whether selected pPS–hallmark associations were attenuated after adjustment for clinically measured adiposity and insulin resistance, we repeated the corresponding hallmark-level models with further adjustment for either body mass index (BMI) or the triglyceride (TG)-to-high-density lipoprotein cholesterol (HDL-C) ratio (TG/HDL-C), a clinically available marker of insulin resistance (32), in both cohorts. Changes in effect estimates were quantified as percentage attenuation on the log odds ratio (OR) or log hazard ratio (HR) scale, with bootstrap confidence intervals and *P* values estimated for coefficient change (see **Supplementary Note 4**).

To account for multiple testing, statistical significance was assessed using Bonferroni-corrected thresholds separately within each cohort and analysis set, defined as 0.05 divided by the number of corresponding tests. Unadjusted *P* values <0.05 were considered nominally significant. Analyses were performed using R version 4.3.1 for the UK Biobank and Python version 3.10.16 for *All of Us*. This study followed the STrengthening the REporting of Genetic Association Studies (STREGA) guidelines (33).

### Data and Resource Availability

Analyses of UK Biobank data were conducted under application 73958. *All of Us* analyses used de-identified participant data accessed through the *All of Us* Researcher Workbench (Controlled Tier, version 8).

## RESULTS

### Participant Characteristics

The analysis included 449,505 participants from the UK Biobank and 374,973 from *All of Us*. Median age at baseline was 58.0 years (IQR 50.0–63.0) in the UK Biobank and 53.0 years (IQR 37.0–65.0) in *All of Us*; females accounted for 53.9% and 60.2% of participants, respectively. The UK Biobank was predominantly of European ancestry (87.2%), whereas *All of Us* was more diverse, with European-, African-, Admixed American-, East Asian-, and South Asian-ancestry participants accounting for 58.0%, 19.3%, 18.4%, 2.5%, and 1.4%, respectively. Detailed cohort characteristics are provided in **Supplementary Tables 4** and **5**.

### Associations of pPSs with Cardiometabolic Traits

In both cohorts, the oPS and all pPSs were positively associated with glycemic traits, while the pPSs showed cluster-specific cardiometabolic patterns, particularly for adiposity, lipid, and liver-related traits, broadly consistent with the original GWAS clusters (**Fig. 1** and **Supplementary Tables 6** and **7**) (2). The beta-cell pPSs were associated with lower adiposity and lower triglycerides, whereas the obesity pPS was associated with greater overall and central adiposity. The lipodystrophy pPS showed a profile characterized by lower BMI, higher waist-to-hip ratio and liver fat percentage, higher TG, and lower HDL-C levels. The body fat and metabolic syndrome pPSs showed weaker but directionally expected profiles, with higher adiposity measures for body fat and a higher-TG, lower-HDL-C pattern for metabolic syndrome. The liver and lipid metabolism pPS was associated with higher liver enzymes and lower low-density lipoprotein cholesterol.

**Figure 1.**
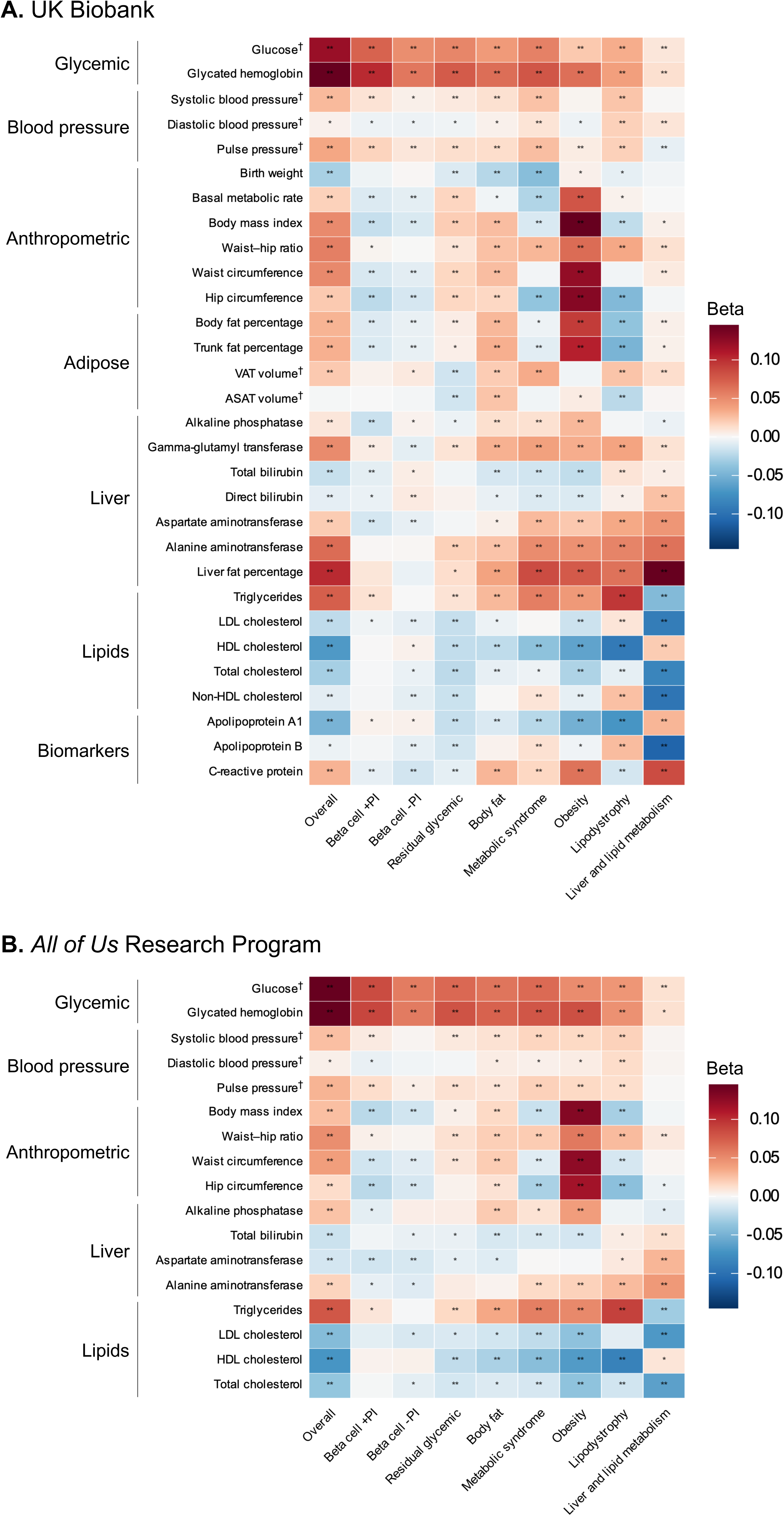
Associations of the overall type 2 diabetes polygenic score (oPS) and partitioned polygenic scores (pPSs) with cardiometabolic traits in the UK Biobank (A) and the *All of Us* Research Program (B). Colors indicate beta coefficients for standardized cardiometabolic traits per 1-SD increase in each standardized oPS or pPS, estimated from linear regression models adjusted for age, sex, and the top 10 genetic principal components; UK Biobank analyses were further adjusted for assessment center and genotyping array. ** indicates Bonferroni-corrected significance, defined as *P* <0.05/270 in the UK Biobank and *P* <0.05/153 in *All of Us*; * indicates unadjusted *P* <0.05; † indicates body mass index-adjusted traits. **Abbreviations**: ASAT, abdominal subcutaneous adipose tissue; HDL, high-density lipoprotein; LDL, low-density lipoprotein; VAT, visceral adipose tissue.

### Associations of PSs with Hallmark-Specific Age-Related Diseases in the UK Biobank

In the UK Biobank, incident analyses over a median follow-up of 14.1 years showed broad positive associations for the oPS and distinct cluster-specific profiles for the pPSs (**Fig. 2** and **Supplementary Table 8**). The oPS was associated with higher risk of incident hallmark-level disease outcomes for most hallmarks, although effect sizes were modest. Across hallmarks, HR point estimates ranged from 1.009 to 1.051 for the first incident disease, from 1.005 to 1.071 for the second incident disease, and from 0.992 to 1.088 for the third incident disease. Among the pPSs, the obesity pPS showed the clearest and most consistent positive associations, with corresponding HR point estimates of 1.002–1.051, 1.013–1.072, and 1.032–1.085. For example, for the third incident telomere attrition-related disease, the HR was 1.065 (95% CI 1.045–1.086; *P* = 8.75 × 10^−11^) for the oPS and 1.085 (1.064–1.105; *P* = 5.30 × 10^−17^) for the obesity pPS. Residual glycemic, body fat, metabolic syndrome, and lipodystrophy pPSs showed smaller and more hallmark-dependent positive associations, whereas both beta-cell pPSs remained close to the null. The liver and lipid metabolism pPS showed mostly null to modest inverse estimates. Tests based on Schoenfeld residuals for the oPS and pPS terms did not indicate non-proportional hazards after Bonferroni correction, although global tests were frequently significant. The findings were similar in Fine-Gray competing-risk models (**Supplementary Fig. 1** and **Supplementary Table 9**).

**Figure 2.**
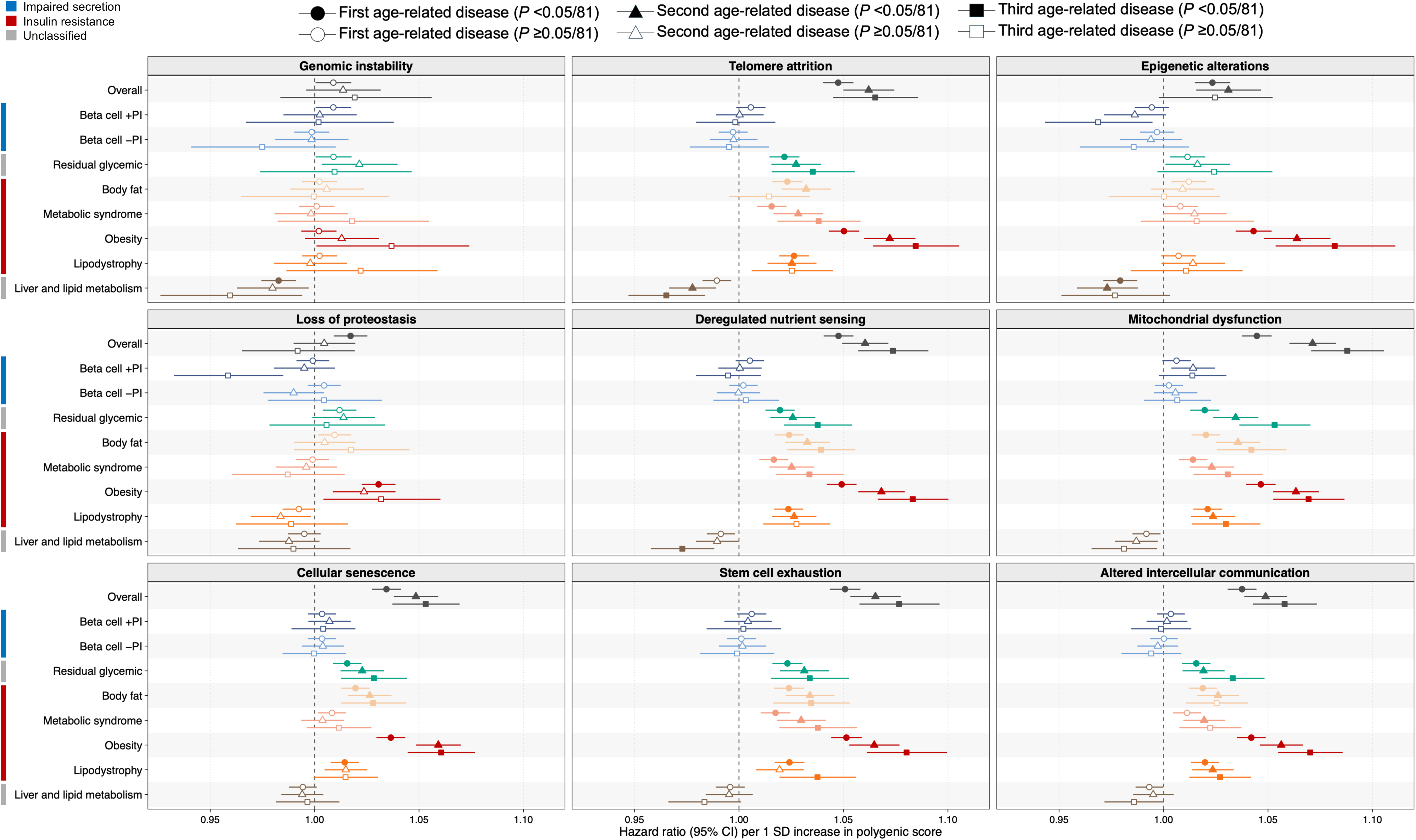
Associations of the overall type 2 diabetes polygenic score (oPS) and partitioned polygenic scores (pPSs) with incident hallmark-specific age-related disease outcomes in the UK Biobank. Incident outcomes were defined as the first, second, or third post-baseline occurrence of an age-related disease within each hallmark, represented by circles, triangles, and squares, respectively. Associations were estimated using Cox proportional hazards models adjusted for age at baseline, sex, assessment center, genotyping array, and the top 10 genetic principal components. Hazard ratios are shown per 1-SD increase in each standardized oPS or pPS. Points show effect estimates and horizontal lines show 95% confidence intervals. Filled symbols indicate Bonferroni-corrected significance, defined as *P* <0.05/81 for 9 scores × 9 hallmark outcomes within each outcome definition; open symbols indicate non-significant associations. Colored side bars indicate the insulin-related profile of each pPS.

### Associations of PSs with Hallmark-Specific Age-Related Diseases in *All of Us*

In *All of Us*, prevalent analyses showed broad positive associations for the oPS and distinct cluster-specific patterns for the pPSs, largely consistent with the UK Biobank incident analyses (**Fig. 3** and **Supplementary Table 10**). Each 1-SD higher standardized oPS was associated with higher odds of at least one disease across all nine hallmarks and with higher odds of within-hallmark multimorbidity for most hallmarks. Among the pPSs, the obesity pPS showed the strongest and most consistent positive associations. Across hallmarks, OR point estimates for the obesity pPS ranged from 1.036 to 1.067, 1.035 to 1.108, and 1.057 to 1.160 across the three prevalent outcome definitions. More modest positive associations were observed for the residual glycemic, body fat, and lipodystrophy pPSs, whereas the metabolic syndrome pPS showed weaker and less consistent associations. For example, for ≥3 co-occurring telomere attrition-related diseases, the OR was 1.095 (95% CI 1.070–1.121; *P* = 3.80 × 10^−14^) for the oPS and 1.116 (1.090–1.142; *P* = 5.43 × 10^−20^) for the obesity pPS, compared with point estimates ranging from 1.031 to 1.043 for the residual glycemic, body fat, and lipodystrophy pPSs. In contrast, the beta-cell pPSs remained close to the null, and the liver and lipid metabolism pPS showed no consistent positive association.

**Figure 3.**
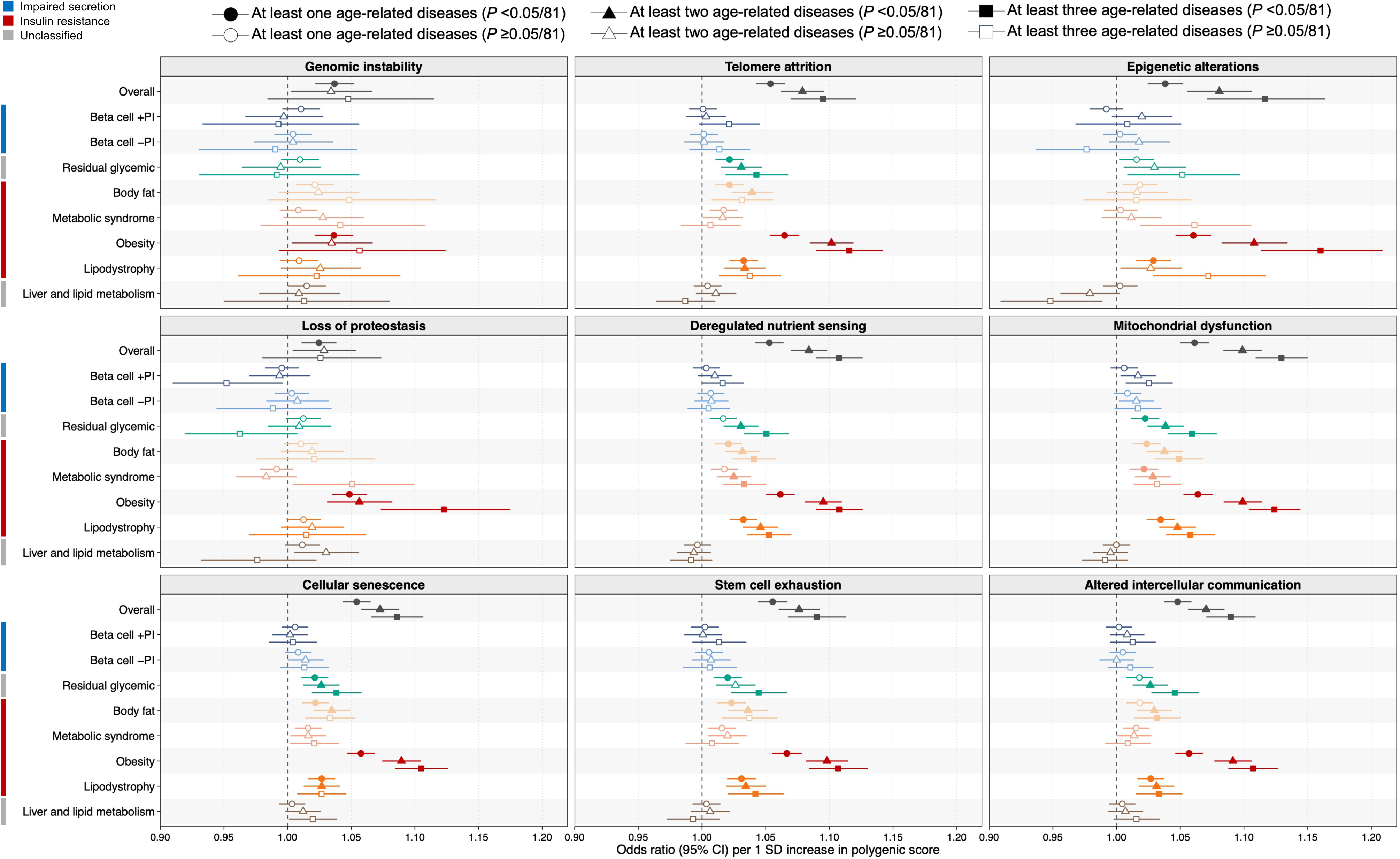
Associations of the overall type 2 diabetes polygenic score (oPS) and partitioned polygenic scores (pPSs) with prevalent hallmark-specific age-related disease outcomes in the *All of Us* Research Program. Prevalent outcomes were defined as the presence of at least one, at least two, or at least three age-related diseases within each hallmark at baseline, represented by circles, triangles, and squares, respectively. Associations were estimated using logistic regression models adjusted for age at baseline, sex, and the top 10 genetic principal components. Odds ratios are shown per 1-SD increase in each standardized oPS or pPS. Points show effect estimates and horizontal lines show 95% confidence intervals. Filled symbols indicate Bonferroni-corrected significance, defined as *P* <0.05/81 for 9 scores × 9 hallmark outcomes within each outcome definition; open symbols indicate non-significant associations. Colored side bars indicate the insulin-related profile of each pPS.

### Associations of PSs with Individual Age-Related Diseases

Individual-disease analyses showed a similar pattern to the hallmark-level analyses, with the broadest positive profiles for the oPS and obesity pPS (**Supplementary Figs. 2** and **3** and **Supplementary Tables 11** and **12**). In the UK Biobank incident analyses, the oPS and obesity pPS were positively associated with 13 and 17 diseases, respectively, after Bonferroni correction. In *All of Us* prevalent analyses, the corresponding numbers were 16 and 21 diseases. Other pPSs showed fewer positive associations, with limited evidence for the beta-cell pPSs and mixed directions for the liver and lipid metabolism pPS.

### Ancestry-Stratified Sensitivity Analyses

Ancestry-stratified sensitivity analyses were broadly consistent with the overall analyses (**Supplementary Tables 13** and **14**). In the UK Biobank, pooled estimates remained most consistently positive for the obesity pPS, whereas the beta-cell pPSs were largely null. Estimates in non-European ancestry strata were less precise, consistent with smaller sample sizes. In *All of Us*, the obesity pPS showed positive pooled estimates across all nine prevalent hallmark outcomes, with pooled ORs ranging from 1.038 (95% CI 1.019–1.059) for genomic instability to 1.064 (1.052–1.076) for stem cell exhaustion. Point estimates for the obesity pPS were consistently positive in European-, African-, and Admixed American-ancestry participants, the three largest ancestry strata, and were also positive but less precise in South Asian- and East Asian-ancestry participants. Other pPSs showed smaller or less consistent estimates, whereas the beta-cell pPSs remained largely null. Between-ancestry heterogeneity was limited overall.

### BMI and TG/HDL-C Adjustment Analyses

The residual glycemic, body fat, metabolic syndrome, obesity, and lipodystrophy pPSs tended to show positive associations with hallmark-level outcomes in the preceding analyses and correspond to clusters characterized by adiposity, insulin resistance, or related cardiometabolic profiles. Therefore, we evaluated the extent to which measured BMI and TG/HDL-C accounted for these associations using BMI and TG/HDL-C adjustment analyses.

BMI adjustment produced the clearest attenuation for the obesity pPS. Across hallmarks, obesity pPS estimates were attenuated by 51.6–74.4% in the UK Biobank and by 6.6–46.1% in All of Us after BMI adjustment, with statistically significant coefficient attenuation for most hallmark outcomes, whereas TG/HDL-C adjustment produced smaller attenuation of 6.6–15.3% and 13.0–19.3%, respectively. In contrast, TG/HDL-C adjustment more strongly attenuated the metabolic syndrome and lipodystrophy pPS estimates, with median attenuation of 53.0% and 63.2% in the UK Biobank and 46.0% and 49.4% in *All of Us*, respectively, and attenuation was statistically significant for most corresponding hallmark outcomes. Changes for the residual glycemic and body fat pPSs were more modest or less consistent. Overall, obesity pPS associations were most strongly reduced after BMI adjustment, whereas metabolic syndrome and lipodystrophy pPS associations were most strongly reduced after TG/HDL-C adjustment (**Figs. 4** and **5** and **Supplementary Tables 15** and **16**).

**Figure 4.**
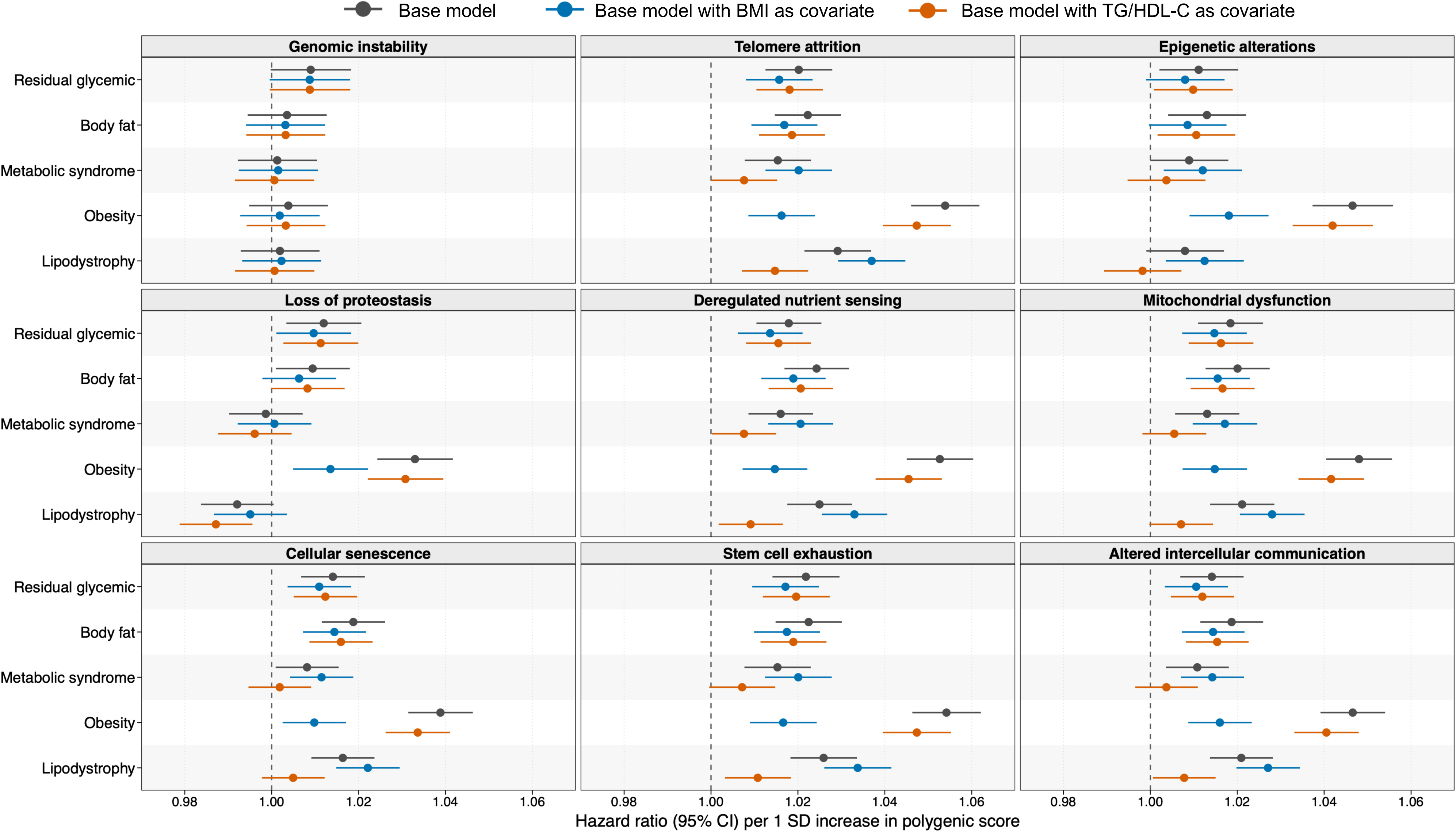
BMI and TG/HDL-C adjustment of partitioned polygenic score (pPS) associations with incident hallmark-specific age-related disease outcomes in the UK Biobank. Associations are shown for the residual glycemic, body fat, metabolic syndrome, obesity, and lipodystrophy pPSs. Incident outcomes were defined as the first post-baseline occurrence of an age-related disease within each hallmark. Base models were adjusted for age at baseline, sex, assessment center, genotyping array, and the top 10 genetic principal components. Adjusted models added either BMI or TG/HDL-C to the base model. Hazard ratios are shown per 1-SD increase in each standardized pPS. Points show effect estimates and horizontal lines show 95% confidence intervals.

**Figure 5.**
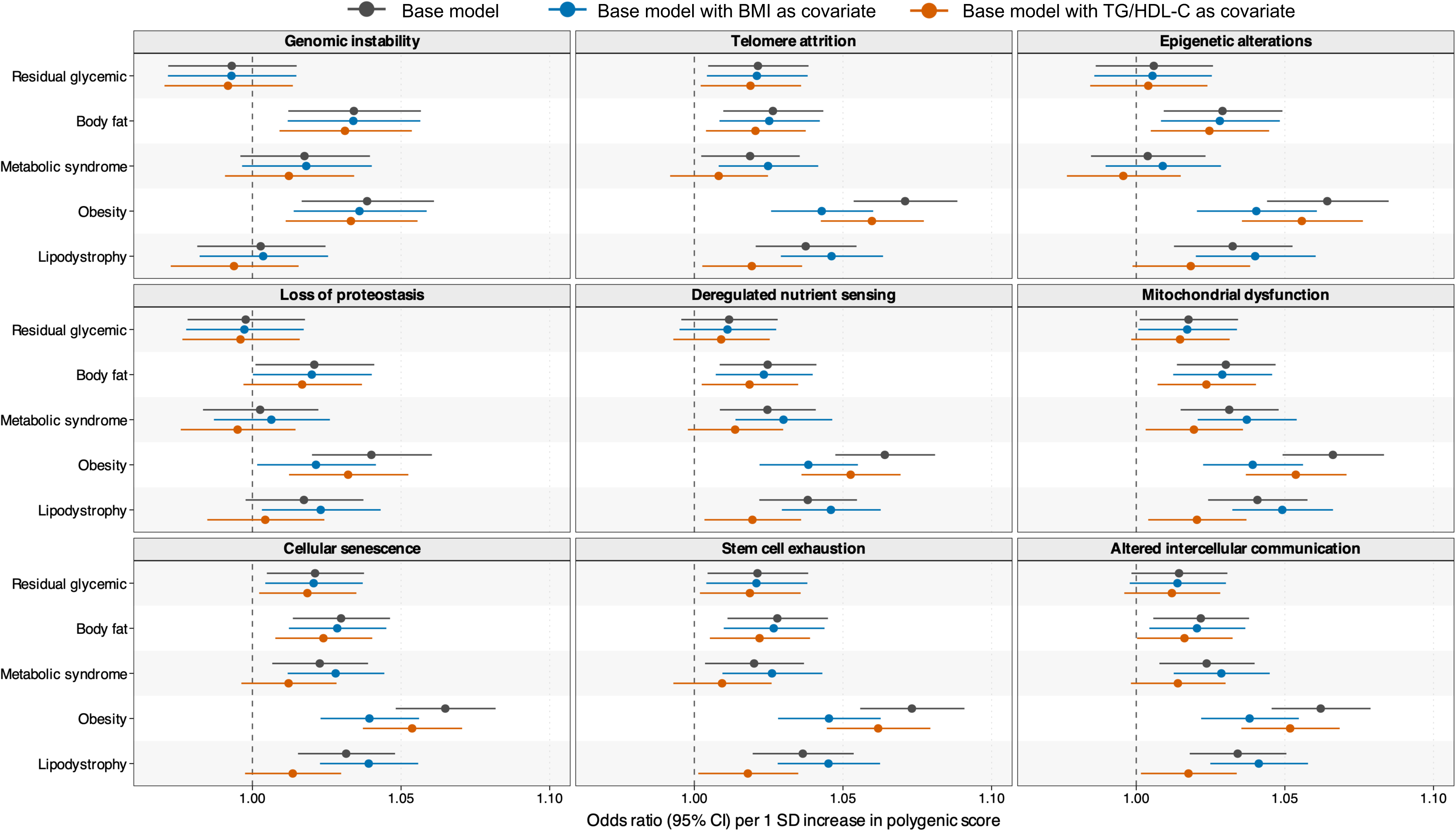
BMI and TG/HDL-C adjustment of partitioned polygenic score (pPS) associations with prevalent hallmark-specific age-related disease outcomes in the *All of Us* Research Program. Associations are shown for the residual glycemic, body fat, metabolic syndrome, obesity, and lipodystrophy pPSs. Prevalent outcomes were defined as the presence of at least one age-related disease within each hallmark at or before baseline. Base models were adjusted for age, sex, and the top 10 genetic principal components. Adjusted models added either BMI or TG/HDL-C to the base model. Odds ratios are shown per 1-SD increase in each standardized pPS. Points show effect estimates and horizontal lines show 95% confidence intervals.

## DISCUSSION

In this study, the oPS showed broad associations with age-related morbidity across hallmarks of aging, whereas T2D pPSs showed heterogeneous associations. Among the pPSs, the obesity pPS showed the strongest positive associations with hallmark-specific outcomes and within-hallmark multimorbidity, and these associations were significantly attenuated after BMI adjustment. Lipodystrophy, metabolic syndrome, body fat, and residual glycemic pPSs showed more modest positive associations, whereas beta-cell pPS associations were close to null. Among the modestly associated pPSs, lipodystrophy and metabolic syndrome associations were significantly attenuated after TG/HDL-C adjustment across most hallmark outcomes. These results suggest that hallmark-defined age-related morbidity linked to T2D genetic susceptibility differs by diabetogenic mechanism and that targeting BMI and TG/HDL-C, both modifiable, may mitigate age-related morbidity conferred by T2D genetic susceptibility across these hallmarks.

Previous studies of T2D pPSs have shown mechanism-specific associations with cardiometabolic profiles, age at T2D onset, disease progression, and diabetes-related complications, including vascular and renal outcomes (2, 8–10). Beyond these T2D-related phenotypes, mechanism-specific associations have also been reported for colorectal cancer and neurological outcomes (11, 12), as well as for circulating metabolite profiles (34, 35). Mendelian randomization studies have also suggested mechanism-specific heterogeneity across non-cardiovascular comorbidities (36, 37). However, whether this heterogeneity extends to broader patterns of clinically manifest age-related morbidity has remained unclear. By grouping age-related diseases according to the hallmarks of aging (14, 17), our study extends this literature from individual traits and selected diseases to a broader, biologically organized spectrum of age-related morbidity.

The positive associations of the obesity pPS across hallmark outcomes align with a previous observational study using the same hallmark framework, in which obesity was associated with higher risk of age-related disease development across all hallmarks (18). Mechanistic and genetic studies also link obesity to multiple biological processes implicated in aging, including mitochondrial dysfunction, cellular senescence, and epigenetic aging (38–40). In our analyses, these associations were reduced after conditioning on BMI, suggesting that measured BMI accounts for a substantial component of obesity pPS-associated hallmark-defined morbidity. A previous study similarly showed that disease risks conferred by BMI-associated variants are largely mediated through measured BMI across several chronic diseases (41); our findings suggest that this BMI-mediated burden is also relevant to broad age-related diseases grouped by the hallmarks of aging. These results support adiposity, indexed clinically by BMI, as a modifiable target for mitigating broader age-related morbidity associated with T2D genetic susceptibility.

Beyond the obesity pPS, lipodystrophy, metabolic syndrome, body fat, and residual glycemic pPSs were positively associated with hallmark-specific outcomes, although less consistently. Their cardiometabolic profiles, together with that of the obesity pPS, are generally aligned with lower insulin sensitivity, although the residual glycemic pPS also reflects impaired insulin secretion (2). These findings suggest that insulin-resistance-related mechanisms may be shared contributors to age-related morbidity across several components of T2D genetic heterogeneity. Insulin resistance, particularly when accompanied by hyperinsulinemia, may promote age-related pathology through sustained insulin/IGF-1 signalling and mTOR activation, processes implicated in longevity control and cellular senescence (21, 22). In the present study, adjustment for TG/HDL-C significantly reduced the associations of the lipodystrophy and metabolic syndrome pPSs with most hallmark-specific outcomes, suggesting that these insulin-resistance-related associations are partly captured by TG/HDL-C. Taken together, BMI and TG/HDL-C are clinically accessible measures that index adiposity and insulin resistance, mechanisms potentially modifiable through lifestyle and therapeutic interventions (42–44). Nonetheless, the stronger associations of the obesity pPS with hallmark-specific outcomes likely reflect the effects of greater adiposity itself in addition to insulin resistance, including hemodynamic burden, mechanical stress, and obesity-related inflammation (38, 45, 46).

The beta-cell pPSs were characterized by impaired insulin secretion without the reduced insulin sensitivity that defined the insulin-resistance-related pPSs discussed above (2). In our study, they also lacked the adverse adiposity- or inflammation-related profile characteristic of the obesity pPS and were generally associated with lower BMI or related body-size measures (**Fig. 1**). This relative lack of accompanying systemic metabolic burden may help explain why their hallmark-level associations were weaker and less consistent. However, these findings should not be interpreted as evidence that impaired insulin secretion or hyperglycemia has limited relevance to aging, because pPSs index inherited susceptibility rather than clinical glycemic exposure or disease course.

The liver and lipid metabolism pPS also requires cautious interpretation. This cluster comprises only three variants and is characterized by higher liver fat and liver-related biomarkers but lower atherogenic lipoprotein measures (2). Its weak or inverse hallmark-level associations may therefore reflect variant-specific pleiotropy and offsetting disease-level effects, rather than a single broadly acting liver- or lipid-related pathway.

An important consideration is that hallmark-specific outcomes were clinically manifest, often overlapping age-related disease groups rather than direct cellular measures. This disease-to-hallmark classification builds on large-scale EHR analyses that identified diseases with age-increasing incidence and mapped them to hallmark domains using literature mining, shared genetic associations, gene-set enrichment, and disease co-occurrence analyses (14, 17). Previous multicohort studies, including UK Biobank, have also shown within-hallmark disease clustering and associations with aging-related exposures (18, 19). These lines of evidence support the use of hallmark-specific disease outcomes as clinical measures of hallmark-organized age-related morbidity in population-based analyses.

Our study has several limitations. First, the cluster definitions were based on hard clustering, assigning each variant to a single cluster despite possible pleiotropy, and the number of variants differed substantially across clusters, affecting precision and interpretability. Second, although the pPSs were derived from multi-ancestry GWAS data, participants of European ancestry still predominated, particularly in the UK Biobank. *All of Us* provided a more diverse comparison cohort with broadly concordant patterns, but smaller ancestry strata yielded heterogeneous or imprecise estimates in ancestry-stratified analyses. Third, although we used UK Biobank-excluded summary statistics for score weights to minimize bias from sample overlap, we retained the index variants and cluster assignments from the source GWAS to preserve the established mechanistic framework and comparability with the original clusters. Variant selection and cluster assignment might therefore have differed if both had been repeated in UK Biobank-excluded data. However, the resulting scores broadly recapitulated the expected cluster-specific cardiometabolic profiles in both cohorts (**Fig. 1**), supporting their use as proxies for these mechanistic clusters. Fourth, outcome ascertainment relied on inpatient records, which may have undercaptured outpatient-managed chronic conditions or delayed apparent onset, although this approach likely improved specificity and robustness of case definition (47).

In conclusion, pPSs of adiposity- and insulin-resistance-related diabetogenic mechanisms were strongly linked to age-related morbidity across hallmarks of aging, whereas pPSs of beta-cell dysfunction were close to null across hallmarks, indicating that the genetic burden of T2D on hallmarks of aging is substantially attributable to adiposity and insulin resistance. Indexed by BMI and TG/HDL-C, adiposity and insulin resistance may act as modifiable factors in the T2D genetic burden, highlighting actionable opportunities to reduce morbidity across hallmarks.

## Data Availability

UK Biobank data are available to approved researchers through the UK Biobank access procedures; analyses in this study were conducted under application 73958. All of Us data are available to registered researchers through the All of Us Researcher Workbench; analyses in this study used de-identified participant data from the Controlled Tier, version 8. Individual-level data cannot be shared by the authors and must be accessed through the respective data access procedures. Summary results generated in this study are provided in the manuscript and supplementary materials.

## Acknowledgments

We thank the participants and staff of the UK Biobank and the *All of Us* Research Program for making this research possible. We thank the Type 2 Diabetes Global Genomics Initiative members for the constructive feedback on the manuscript. M.H. is supported by the Japan Student Services Organization (JASSO; Graduate scholarship for degree-seeking study abroad selected under the special priority framework for doctoral studies at top-tier global universities in STEM fields) and the Watanabe Foundation (6th Toshizo Watanabe International Scholarship). C.N.S. was supported by the American Diabetes Association (11-22-JDFPM-06). T.L. is supported by start-up funding from the Office of the Vice Chancellor for Research and Graduate Education, the School of Medicine and Public Health, and the Department of Population Health Sciences at the University of Wisconsin–Madison.

## Author Contributions

M.H. and S.Y. conceived and designed the study. M.H. performed the analyses and wrote the first draft of the manuscript. C.Z. and C.N.S. contributed data used in the analyses. All authors contributed to the interpretation of results, critically revised the manuscript for important intellectual content, and approved the final version of the manuscript. S.Y. is the guarantor of this work and, as such, had full access to all the data in the study and takes responsibility for the integrity of the data and the accuracy of the data analysis.

## References

1. Leslie RD, Ma RCW, Franks PW, Nadeau KJ, Pearson ER, Redondo MJ. Understanding diabetes heterogeneity: key steps towards precision medicine in diabetes. Lancet Diabetes Endocrinol. 2023;11(11):848–60.

2. Suzuki K, Hatzikotoulas K, Southam L, Taylor HJ, Yin X, Lorenz KM, et al. Genetic drivers of heterogeneity in type 2 diabetes pathophysiology. Nature. 2024;627(8003):347–57.

3. Smith K, Deutsch AJ, McGrail C, Kim H, Hsu S, Huerta-Chagoya A, et al. Multi-ancestry polygenic mechanisms of type 2 diabetes. Nat Med. 2024;30(4):1065–74.

4. Ahlqvist E, Storm P, Karajamaki A, Martinell M, Dorkhan M, Carlsson A, et al. Novel subgroups of adult-onset diabetes and their association with outcomes: a data-driven cluster analysis of six variables. Lancet Diabetes Endocrinol. 2018;6(5):361–9.

5. Kalyani RR, Golden SH, Cefalu WT. Diabetes and Aging: Unique Considerations and Goals of Care. Diabetes Care. 2017;40(4):440–3.

6. Harding JL, Pavkov ME, Magliano DJ, Shaw JE, Gregg EW. Global trends in diabetes complications: a review of current evidence. Diabetologia. 2019;62(1):3–16.

7. Bonnefond A, Florez JC, Loos RJF, Froguel P. Dissection of type 2 diabetes: a genetic perspective. Lancet Diabetes Endocrinol. 2025;13(2):149–64.

8. Reynolds KM, Sha D, Sun Q, Parsa A, Chen J, Rincon-Choles H, et al. Diabetes Genetic Clusters and Clinical Outcomes in the Chronic Renal Insufficiency Cohort. Clin J Am Soc Nephrol. 2026;21(2):221–8.

9. Yu G, Tam CHT, Lim CKP, Shi M, Lau ESH, Ozaki R, et al. Type 2 diabetes pathway-specific polygenic risk scores elucidate heterogeneity in clinical presentation, disease progression and diabetic complications in 18,217 Chinese individuals with type 2 diabetes. Diabetologia. 2025;68(3):602–14.

10. Reynolds KM, Sun Q, Zhang Y, Umans J, Cole SA, Morris AP, et al. Diabetes Genetic Clusters and Clinical Outcomes in American Indians. Diabetes. 2025;74(11):2132–9.

11. Zhou X, Sevilla-Gonzalez M, Phipps AI, Udler M, Castellvi-Bel S, Chan AT, et al. Diabetogenic processes for insulin resistance-linked hyperinsulinaemia are associated with colorectal cancer. Diabetologia. 2026;69(4):942–52.

12. Sarnowski C, Zhang Y, Ammous F, Shade LMP, DiCorpo D, Jian X, et al. Association of genetic scores related to insulin resistance with neurological outcomes in ancestrally diverse cohorts from the Trans-Omics for Precision Medicine (TOPMed) program. Commun Biol. 2025;8(1):1352.

13. Kuan V, Denaxas S, Gonzalez-Izquierdo A, Direk K, Bhatti O, Husain S, et al. A chronological map of 308 physical and mental health conditions from 4 million individuals in the English National Health Service. Lancet Digit Health. 2019;1(2):e63–e77.

14. Fraser HC, Kuan V, Johnen R, Zwierzyna M, Hingorani AD, Beyer A, et al. Biological mechanisms of aging predict age-related disease co-occurrence in patients. Aging Cell. 2022;21(4):e13524.

15. Lopez-Otin C, Blasco MA, Partridge L, Serrano M, Kroemer G. The hallmarks of aging. Cell. 2013;153(6):1194–217.

16. Lopez-Otin C, Blasco MA, Partridge L, Serrano M, Kroemer G. Hallmarks of aging: An expanding universe. Cell. 2023;186(2):243–78.

17. Kuan V, Fraser HC, Hingorani M, Denaxas S, Gonzalez-Izquierdo A, Direk K, et al. Data-driven identification of ageing-related diseases from electronic health records. Sci Rep. 2021;11(1):2938.

18. Kivimaki M, Frank P, Pentti J, Xu X, Vahtera J, Ervasti J, et al. Obesity and risk of diseases associated with hallmarks of cellular ageing: a multicohort study. Lancet Healthy Longev. 2024;5(7):e454–e63.

19. Kivimaki M, Pentti J, Frank P, Liu F, Blake A, Nyberg ST, et al. Social disadvantage accelerates aging. Nat Med. 2025;31(5):1635–43.

20. Tuduri E, Soriano S, Almagro L, Montanya E, Alonso-Magdalena P, Nadal A, et al. The pancreatic beta-cell in ageing: Implications in age-related diabetes. Ageing Res Rev. 2022;80:101674.

21. Lopez-Otin C, Galluzzi L, Freije JMP, Madeo F, Kroemer G. Metabolic Control of Longevity. Cell. 2016;166(4):802–21.

22. Spinelli R, Baboota RK, Gogg S, Beguinot F, Bluher M, Nerstedt A, et al. Increased cell senescence in human metabolic disorders. J Clin Invest. 2023;133(12).

23. Sudlow C, Gallacher J, Allen N, Beral V, Burton P, Danesh J, et al. UK biobank: an open access resource for identifying the causes of a wide range of complex diseases of middle and old age. PLoS Med. 2015;12(3):e1001779.

24. Bycroft C, Freeman C, Petkova D, Band G, Elliott LT, Sharp K, et al. The UK Biobank resource with deep phenotyping and genomic data. Nature. 2018;562(7726):203–9.

25. All of Us Research Program I, Denny JC, Rutter JL, Goldstein DB, Philippakis A, Smoller JW, et al. The “All of Us” Research Program. N Engl J Med. 2019;381(7):668–76.

26. All of Us Research Program Genomics I. Genomic data in the All of Us Research Program. Nature. 2024;627(8003):340–6.

27. Chang CC, Chow CC, Tellier LC, Vattikuti S, Purcell SM, Lee JJ. Second-generation PLINK: rising to the challenge of larger and richer datasets. Gigascience. 2015;4:7.

28. Klann JG, Joss MAH, Embree K, Murphy SN. Data model harmonization for the All Of Us Research Program: Transforming i2b2 data into the OMOP common data model. PLoS One. 2019;14(2):e0212463.

29. Austin PC, Fine JP. Practical recommendations for reporting Fine-Gray model analyses for competing risk data. Stat Med. 2017;36(27):4391–400.

30. Ding Y, Hou K, Xu Z, Pimplaskar A, Petter E, Boulier K, et al. Polygenic scoring accuracy varies across the genetic ancestry continuum. Nature. 2023;618(7966):774–81.

31. DerSimonian R, Laird N. Meta-analysis in clinical trials revisited. Contemp Clin Trials. 2015;45(Pt A):139–45.

32. McLaughlin T, Abbasi F, Cheal K, Chu J, Lamendola C, Reaven G. Use of metabolic markers to identify overweight individuals who are insulin resistant. Ann Intern Med. 2003;139(10):802–9.

33. Little J, Higgins JP, Ioannidis JP, Moher D, Gagnon F, von Elm E, et al. STrengthening the REporting of Genetic Association Studies (STREGA): an extension of the STROBE statement. PLoS Med. 2009;6(2):e22.

34. Peng R, Liu L, Chu X, Xia Z, Fu Q, Silva LF, et al. Plasma metabolite association profiles for type 2 diabetes genetic clusters in Finnish men. Diabetologia. 2026.

35. Bragg F, Alegre-Diaz J, Trichia E, Torres JM, Baca P, Garcilazo-Avila A, et al. Metabolomic Profile of Genetic Liability to Type 2 Diabetes Among 125,000 Mexican Adults: A Mendelian Randomization Study. Diabetes Care. 2026.

36. Arruda AL, Bocher O, Taylor HJ, Cammann D, Yoshiji S, Yin X, et al. The effect of type 2 diabetes genetic predisposition on non-cardiovascular comorbidities. Nat Commun. 2025;16(1):9042.

37. Bala R, Handley D, Gillett A, Green H, Bowden J, Wood A, et al. Evidence of bidirectional relationship between type 2 diabetes and depression; a Mendelian randomization study. Mol Psychiatry. 2025;30(11):5013–23.

38. Tam BT, Morais JA, Santosa S. Obesity and ageing: Two sides of the same coin. Obes Rev. 2020;21(4):e12991.

39. Franceschi C. Healthy ageing in 2016: Obesity in geroscience - is cellular senescence the culprit? Nat Rev Endocrinol. 2017;13(2):76–8.

40. Li J, Wang W, Yang Z, Qiu L, Ren Y, Wang D, et al. Causal association of obesity with epigenetic aging and telomere length: a bidirectional mendelian randomization study. Lipids Health Dis. 2024;23(1):78.

41. Einarsson G, Thorleifsson G, Steinthorsdottir V, Zink F, Helgason H, Olafsdottir T, et al. Sequence variants associated with BMI affect disease risk through BMI itself. Nat Commun. 2024;15(1):9335.

42. Kim MS, Shim I, Fahed AC, Do R, Park WY, Natarajan P, et al. Association of genetic risk, lifestyle, and their interaction with obesity and obesity-related morbidities. Cell Metab. 2024;36(7):1494–503 e3.

43. Pollin TI, Isakova T, Jablonski KA, de Bakker PI, Taylor A, McAteer J, et al. Genetic modulation of lipid profiles following lifestyle modification or metformin treatment: the Diabetes Prevention Program. PLoS Genet. 2012;8(8):e1002895.

44. German J, Cordioli M, Tozzo V, Urbut S, Arumae K, Smit RAJ, et al. Association between plausible genetic factors and weight loss from GLP1-RA and bariatric surgery. Nat Med. 2025;31(7):2269–76.

45. Heymsfield SB, Wadden TA. Mechanisms, Pathophysiology, and Management of Obesity. N Engl J Med. 2017;376(3):254–66.

46. Andonian BJ, Hippensteel JA, Abuabara K, Boyle EM, Colbert JF, Devinney MJ, et al. Inflammation and aging-related disease: A transdisciplinary inflammaging framework. Geroscience. 2025;47(1):515–42.

47. MacRae C, Morales D, Mercer SW, Lone N, Lawson A, Jefferson E, et al. Impact of data source choice on multimorbidity measurement: a comparison study of 2.3 million individuals in the Welsh National Health Service. BMC Med. 2023;21(1):309.

