## Supplementary Figure for "Type 2 Diabetes Partitioned Polygenic Scores Are Differentially Associated with Aging Hallmarks"

**Supplementary Figure 1**. Associations of the overall type 2 diabetes polygenic score (oPS) and partitioned polygenic scores (pPSs) with incident hallmark-specific age-related disease outcomes from Fine-Gray competing-risk models in the UK Biobank.

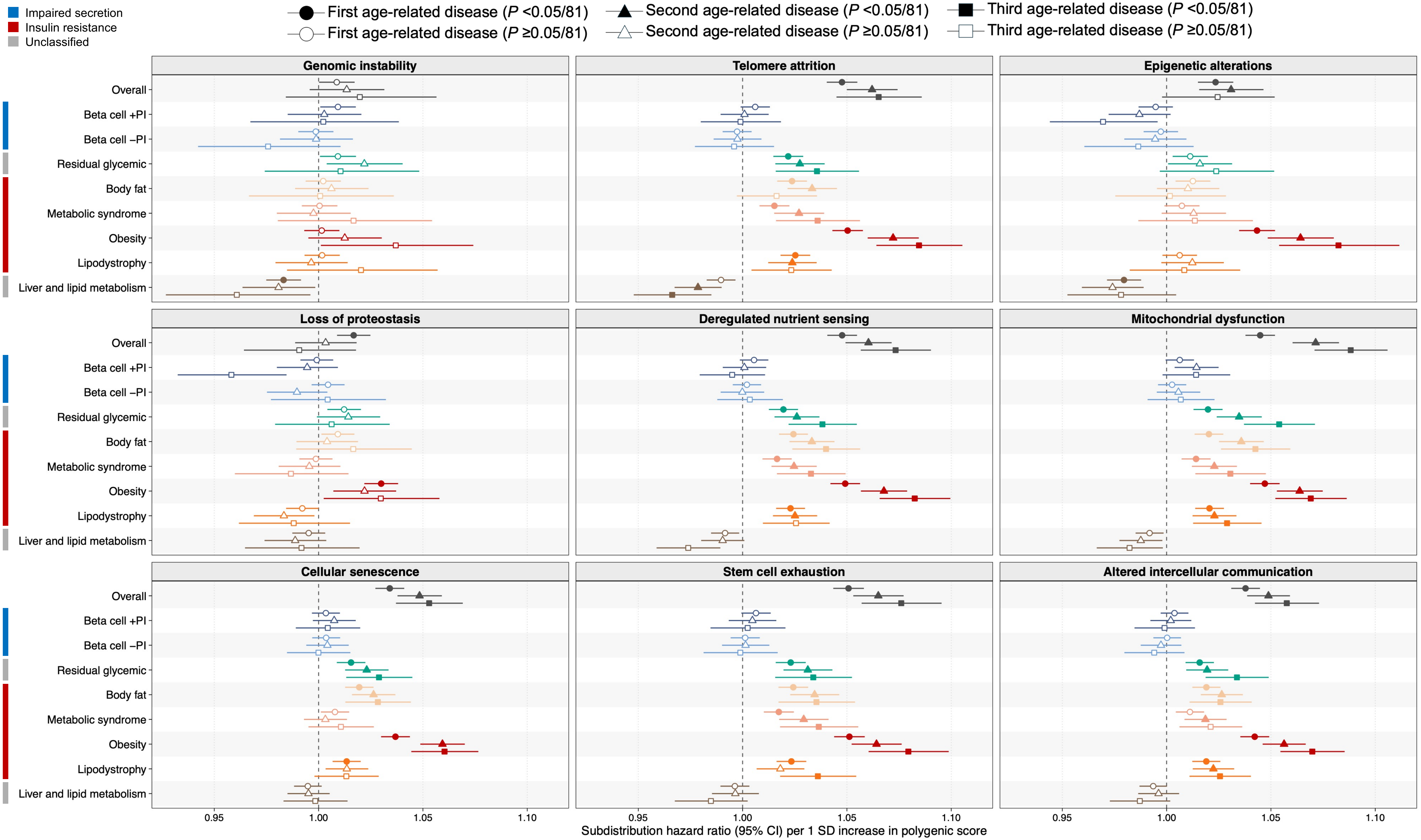


Hallmark-specific outcomes were defined as the first, second, or third post-baseline occurrence of an age-related disease assigned to each hallmark, represented by circles, triangles, and squares, respectively. Associations were estimated using Fine-Gray subdistribution hazard models, treating death not attributable to diseases within the corresponding hallmark as a competing event, and adjusted for age at baseline, sex, assessment center, genotyping array, and the top 10 genetic principal components. Subdistribution hazard ratios are shown per 1-SD increase in each standardized oPS or pPS. Points show effect estimates and horizontal lines show 95% confidence intervals. Filled symbols indicate Bonferroni-corrected significance, defined as $P < 0.05/81$ for 9 scores × 9 hallmark outcomes within each outcome definition; open symbols indicate non-significant associations. Colored side bars indicate the insulin-related profile of each pPS.

**Supplementary Figure 2**. Associations of the overall type 2 diabetes polygenic score (oPS) and partitioned polygenic scores (pPSs) with incident individual age-related diseases in the UK Biobank.

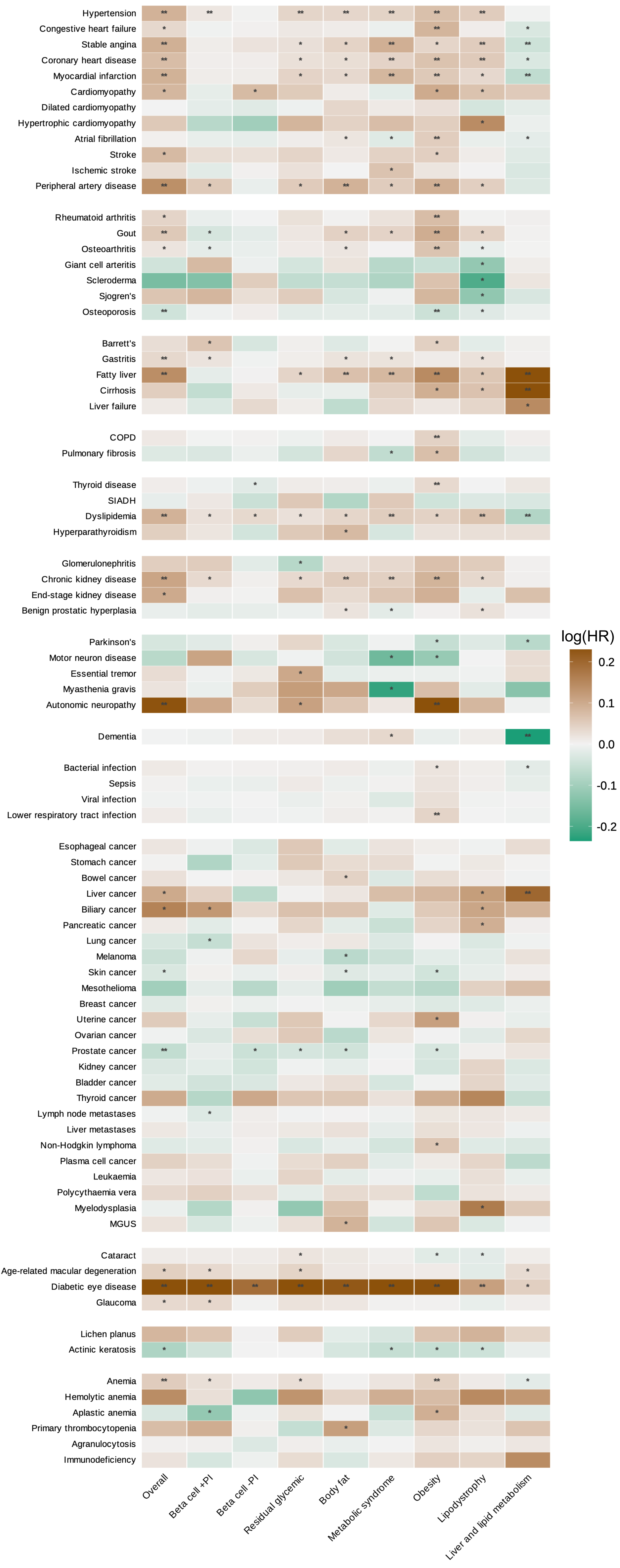


The heatmap shows the log hazard ratios from Cox proportional hazards models for incident age-related diseases occurring after baseline. Models were adjusted for age at baseline, sex, the top 10 genetic principal components, assessment center, and genotyping array. Colors indicate the magnitude and direction of the association as log hazard ratios per 1-SD increase in each standardized oPS or pPS. ** indicates Bonferroni-corrected significance, defined as $P$ <0.05/729 for 9 scores × 81 diseases; * indicates unadjusted $P$ <0.05. **Abbreviations**: COPD, chronic obstructive pulmonary disease; MGUS, monoclonal gammopathy of undetermined significance; PS, polygenic score; SIADH, syndrome of inappropriate antidiuretic hormone secretion.

**Supplementary Figure 3**. Associations of the overall type 2 diabetes polygenic score (oPS) and partitioned polygenic scores (pPSs) with prevalent individual age-related diseases in the *All of Us* Research Program.

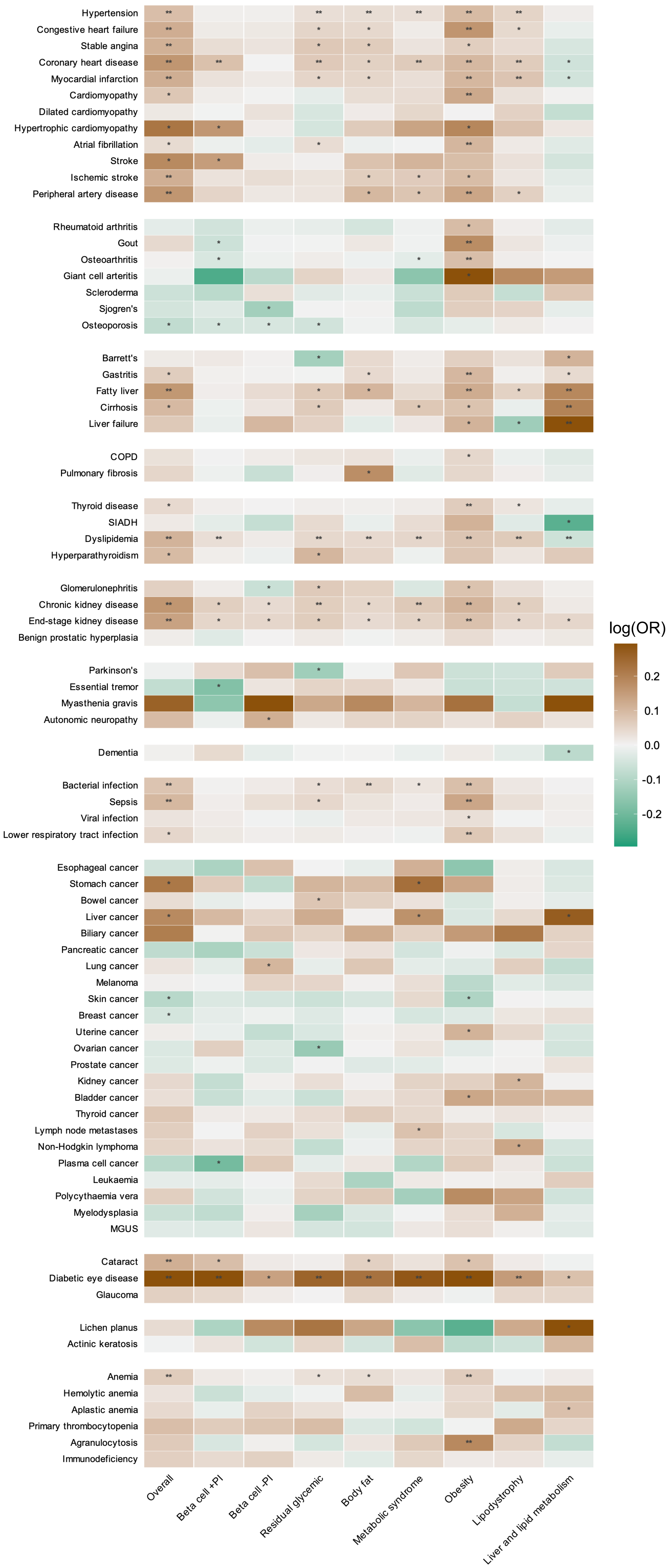


The heatmap shows the log odds ratios from logistic regression models for prevalent age-related diseases defined at or before baseline. Models were adjusted for age at baseline, sex, and the top 10 genetic principal components. Colors indicate the magnitude and direction of the association as log odds ratios per 1-SD increase in each standardized PS or pPS. ** indicates Bonferroni-corrected significance, defined as $P < 0.05/729$ for 9 scores × 81 diseases; * indicates unadjusted $P < 0.05$.

**Abbreviations**: COPD, chronic obstructive pulmonary disease; MGUS, monoclonal gammopathy of undetermined significance; PS, polygenic score; SIADH, syndrome of inappropriate antidiuretic hormone secretion.
