## Supplementary Note for "Type 2 Diabetes Partitioned Polygenic Scores Are Differentially Associated with Aging Hallmarks"

**Supplementary Note 1.** Derivation and implementation of hallmark-specific age-related disease definitions.

Our outcome definitions were based on the hallmark-disease framework described by Kuan et al. (1) and Fraser et al. (2), as well as its subsequent application in multicohort studies using ICD-10-coded data (3, 4). In this framework, age-related diseases are classified according to their putative links to the nine hallmarks of aging (5, 6), enabling analyses at both the individual disease level and the broader hallmark level. The original framework included 83 diseases overall, with 27 to 30 diseases assigned to each hallmark, and some diseases contributing to more than one hallmark; therefore, hallmark-level outcomes were not mutually exclusive. In the present study, diabetes-related disease definitions were excluded because diabetes-related genetic liability was examined as the exposure rather than the outcome.

In the UK Biobank, hallmark assignments were implemented at the level of ICD-10 codes. After excluding diabetes-related disease definitions, 81 age-related diseases were retained for analysis. When both a 3-character ICD-10 code and a more specific 4-character descendant code were assigned to different disease entities, the more specific code was prioritized to avoid double counting of the same clinical event across diseases. For example, C22.1 was assigned to biliary cancer, whereas the broader parent code C22 was used for liver cancer; accordingly, records coded as C22.1 were classified only as biliary cancer and not as liver cancer. In multimorbidity analyses, ICD-10 codes assigned to more than one disease entity (A37, A40, I25.5, J12–J15, and J86) were reassigned to a single clinically more specific disease category so that the same event was not counted more than once within a hallmark. The ICD-10 code definitions applied in the UK Biobank analyses are provided in **Supplementary Table 2**.

In the *All of Us* Research Program, the ICD-10-based definitions were mapped to standard Observational Medical Outcomes Partnership (OMOP) Condition concepts for inpatient diagnoses (7). Because this mapping was not always one-to-one, the resulting concept sets were manually reviewed and refined to preserve consistency with the source definitions and intended disease phenotypes. Concepts judged to be clearly inconsistent with the target phenotype were excluded, and when more than one disease assignment was possible, the concept was assigned to the more clinically specific disease category. The disease definition for liver metastases in the original framework could not be adequately represented using inpatient OMOP Condition concepts and was therefore excluded. Accordingly, 80 age-related diseases were retained for analyses in *All of Us*. The OMOP concept definitions used in the *All of Us* analyses are provided in **Supplementary Table 3**.

**Supplementary Note 2**. Associations of cardiometabolic traits with partitioned polygenic risk scores for type 2 diabetes.

We examined associations of the overall polygenic score (oPS) and partitioned polygenic scores (pPSs) for type 2 diabetes with cardiometabolic traits in the UK Biobank and the *All of Us* Research Program to assess whether each score recapitulated the cardiometabolic patterns reported in the original multi-ancestry genome-wide association study (8). We prioritized cardiometabolic traits included in the original clustering analysis and, in each cohort, analyzed those that were available and could be derived from the available data.

In the UK Biobank, we analyzed 30 cardiometabolic traits derived primarily from baseline measurements (instance 0): glucose (UK Biobank data field 30740), glycated hemoglobin (HbA1c; data field 30750), systolic blood pressure (SBP; data field 4080), diastolic blood pressure (DBP; data field 4079), pulse pressure (PP), birth weight (data field 20022), basal metabolic rate (data field 23105), body mass index (BMI; data field 21001), waist circumference (WC; data field 48), hip circumference (HC; data field 49), waist-hip ratio, body fat percentage (data field 23099), trunk fat percentage (data field 23127), visceral adipose tissue volume (data field 22407), abdominal subcutaneous adipose tissue volume (data field 22408), alkaline phosphatase (ALP; data field 30610), gamma-glutamyl transferase (data field 30730), total bilirubin (data field 30840), direct bilirubin (data field 30660), aspartate aminotransferase (AST; data field 30650), alanine aminotransferase (ALT; data field 30620), liver fat percentage (data field 40061), triglycerides (data field 30870), low-density lipoprotein (LDL) cholesterol (data field 30780), high-density lipoprotein (HDL) cholesterol (data field 30760), total cholesterol (TC; data field 30690), non-HDL cholesterol, apolipoprotein A1 (data field 30630), apolipoprotein B (data field 30640), and C-reactive protein (data field 30710). SBP and DBP were defined as the mean of the two baseline measurements, and PP was calculated as SBP minus DBP. Waist-hip ratio was calculated as WC divided by HC, and non-HDL cholesterol was calculated as TC minus HDL cholesterol. For imaging-derived traits, visceral adipose tissue volume, abdominal subcutaneous adipose tissue volume, and liver fat percentage were taken from imaging instance 2 when available; otherwise, instance 3 was used if available. For these traits, age, BMI, and assessment center (data field 54) were obtained from the corresponding assessment instance.

In *All of Us*, we analyzed 17 cardiometabolic traits derived from program-collected physical measurements and laboratory measurements recorded in the Observational Medical Outcomes Partnership (OMOP) Common Data Model: glucose (concept ID 3004501), HbA1c (3004410), SBP (3004249), DBP (3012888), PP, BMI, WC (40759207), HC (40765148), waist-hip ratio, ALP (3035995), AST (3013721), ALT (3006923), total bilirubin (3024128), triglycerides (3022192), LDL cholesterol (3028288), HDL cholesterol (3007070), and TC (3027114). BMI was calculated as weight in kilograms (3025315) divided by height in meters (3036277) squared, PP as SBP minus DBP, and waist-hip ratio as WC divided by HC. For direct physical measurements, multiple values recorded on the same day were collapsed to their median, and the value from the date closest to the primary consent date was selected for each participant and trait; in *All of Us*, the primary consent date was used as the baseline reference date. For laboratory traits, measurements were restricted to those obtained within ±365 days of the primary consent date, and the median of all eligible values was used for each trait. Prespecified unit restrictions and broad plausibility filters were applied before deriving participant-level trait values.

For association analyses, selected traits were log-transformed and all traits were then standardized to mean 0 and SD 1. In the UK Biobank, log transformation was applied to triglycerides, C-reactive protein, ALP, gamma-glutamyl transferase, AST, ALT, direct bilirubin, total bilirubin, and liver fat percentage; in *All of Us*, log transformation was applied to triglycerides, ALP, AST, ALT, and total bilirubin. Associations were examined using complete-case linear regression. In each cohort, pPSs and the oPS were residualized for the top 10 genetic principal components (PCs) and standardized within that cohort before analyses; the PCs were obtained from UK Biobank data field 22009 and from auxiliary genomic ancestry data provided in *All of Us*. Models were adjusted for age, sex, and the top 10 genetic PCs, with additional adjustment for assessment center and genotyping array (data field 22000) in the UK Biobank. BMI-adjusted models were used for glucose, SBP, DBP, and PP in both cohorts, and additionally for visceral adipose tissue volume and abdominal subcutaneous adipose tissue volume in the UK Biobank. Bonferroni correction was applied separately within each cohort, using thresholds of $P < 0.05/270$ for the UK Biobank analysis and $P < 0.05/153$ for the *All of Us* analysis, corresponding to 9 scores × 30 traits and 9 scores × 17 traits, respectively.

**Supplementary Note 3**. Ancestry-stratified sensitivity analyses.

We performed ancestry-stratified sensitivity analyses to evaluate whether the main associations were directionally consistent across genetic ancestry groups. These analyses focused on the primary hallmark-level outcome in each cohort: incident first hallmark-specific age-related disease in the UK Biobank and prevalent hallmark-specific age-related disease in the *All of Us* Research Program, defined as the presence of at least one disease assigned to each hallmark.

In the UK Biobank, genetic ancestry was defined using the ancestry classification in the UK Biobank data field 30079. Analyses included participants assigned to African, Admixed American, Central/South Asian, East Asian, or European ancestry. In *All of Us*, genetic ancestry was assigned using the auxiliary genomic ancestry classifications, based on genetic similarity to global reference populations. Analyses included participants assigned to African, Admixed American, East Asian, European, or South Asian ancestry. Participants without one of these ancestry assignments were included in the primary analyses but not in ancestry-stratified sensitivity analyses.

For each ancestry group, we used ancestry-specific weights to calculate the overall polygenic score (oPS) and the eight partitioned polygenic scores (pPSs) for type 2 diabetes (T2D). For the European and South Asian strata, weights were derived from genome-wide association study summary statistics excluding UK Biobank participants, consistent with the primary analysis. Scores were calculated in PLINK v2.0 as weighted sums of risk-allele dosages aligned to the T2D risk-increasing allele, using available variants in each cohort (9). Before association analyses, each ancestry-specific PS was standardized within the corresponding ancestry group and cohort.

In the UK Biobank, ancestry-stratified analyses of incident first hallmark-specific diseases were performed using Cox proportional hazards models after excluding participants with the corresponding hallmark-specific disease at baseline. In *All of Us*, ancestry-stratified analyses of prevalent hallmark-specific diseases were performed using logistic regression. Models used the same covariate adjustment as the primary analyses: age, sex, and the top 10 genetic principal components in both cohorts, with additional adjustment for assessment center (data field 54) and genotyping array (data field 22000) in the UK Biobank.

Within each cohort, ancestry-specific association estimates were pooled using DerSimonian-Laird random-effects meta-analysis (10), with hazard ratios pooled for the UK Biobank incident analyses and odds ratios pooled for the *All of Us* prevalent analyses. Between-ancestry heterogeneity was summarized using the $I^2$ statistic and Cochran's Q test.

**Supplementary Note 4**. BMI and TG/HDL-C adjustment analyses.

We assessed whether selected type 2 diabetes (T2D) partitioned polygenic score (pPS) associations with hallmark-level age-related disease outcomes were attenuated after adjustment for measured adiposity and insulin resistance. Body mass index (BMI) was used as a measure of adiposity, and the triglyceride-to–high-density lipoprotein cholesterol ratio (TG/HDL-C), calculated as TG divided by HDL-C, was used as a clinically available marker of insulin resistance (11). Analyses included five pPSs that showed positive associations in at least some hallmark-level analyses or represented adiposity- or insulin resistance–related clusters: residual glycemic, body fat, metabolic syndrome, obesity, and lipodystrophy.

In the UK Biobank, BMI, TG, and HDL-C were obtained from baseline assessment instance 0 using data fields 21001, 30870, and 30760, respectively, and TG/HDL-C was calculated from same-instance TG and HDL-C. In *All of Us*, BMI was calculated from program-collected weight and height measurements corresponding to OMOP concept IDs 3025315 and 3036277. Same-day values were collapsed to the median, and the value closest to the primary consent date was selected. TG and HDL-C were obtained from OMOP laboratory concept IDs 3022192 and 3007070 within ±365 days of the primary consent date. TG/HDL-C was calculated only from same-day TG and HDL-C measurements, after collapsing same-day values to the median and selecting the lipid pair closest to the primary consent date.

Participants were restricted to those with available BMI, TG/HDL-C, covariates, and the corresponding hallmark-level outcome. Within this common analytic sample, we fitted a base model, a BMI-adjusted model, and a TG/HDL-C-adjusted model for each pPS–outcome pair, ensuring that differences in pPS coefficients reflected covariate adjustment rather than differences in sample composition. Incident outcomes in the UK Biobank were analyzed using Cox proportional hazards models, and prevalent outcomes in *All of Us* were analyzed using logistic regression. Base models were adjusted for age, sex, and the top 10 genetic principal components, with additional adjustment for assessment center and genotyping array in the UK Biobank. Adjusted models added BMI or TG/HDL-C to the base model.

Percentage attenuation was calculated on the log hazard ratio scale in the UK Biobank and on the log odds ratio scale in *All of Us* as $100 \times (\beta\_base - \beta\_adjusted) / \beta\_base$, where $\beta\_base$ and $\beta\_adjusted$ denote the pPS coefficients from the base and adjusted models fitted in the same common analytic sample. Positive values indicate smaller estimates after adjustment, whereas negative values indicate larger estimates. Bootstrap resampling was used to estimate 95% confidence intervals for percentage attenuation, and *P* values were calculated for coefficient change, defined as $\beta\_base - \beta\_adjusted$ (12). Statistical significance for coefficient change was assessed using Bonferroni correction across 90 attenuation tests in each cohort, corresponding to five pPSs, two adjustment models, and nine hallmark outcomes: $P < 0.05/90 = 5.56 \times 10^{-4}$.

**Supplementary Note References**


1. Kuan V, Fraser HC, Hingorani M, Denaxas S, Gonzalez-Izquierdo A, Direk K, et al. Data-driven identification of ageing-related diseases from electronic health records. Sci Rep. 2021;11(1):2938.
2. Fraser HC, Kuan V, Johnen R, Zwierzyna M, Hingorani AD, Beyer A, et al. Biological mechanisms of aging predict age-related disease co-occurrence in patients. Aging Cell. 2022;21(4):e13524.
3. Kivimaki M, Pentti J, Frank P, Liu F, Blake A, Nyberg ST, et al. Social disadvantage accelerates aging. Nat Med. 2025;31(5):1635-43.
4. Kivimaki M, Frank P, Pentti J, Xu X, Vahtera J, Ervasti J, et al. Obesity and risk of diseases associated with hallmarks of cellular ageing: a multicohort study. Lancet Healthy Longev. 2024;5(7):e454-e63.
5. Lopez-Otin C, Blasco MA, Partridge L, Serrano M, Kroemer G. The hallmarks of aging. Cell. 2013;153(6):1194-217.
6. Lopez-Otin C, Blasco MA, Partridge L, Serrano M, Kroemer G. Hallmarks of aging: An expanding universe. Cell. 2023;186(2):243-78.
7. Klann JG, Joss MAH, Embree K, Murphy SN. Data model harmonization for the All Of Us Research Program: Transforming i2b2 data into the OMOP common data model. PLoS One. 2019;14(2):e0212463.
8. Suzuki K, Hatzikotoulas K, Southam L, Taylor HJ, Yin X, Lorenz KM, et al. Genetic drivers of heterogeneity in type 2 diabetes pathophysiology. Nature. 2024;627(8003):347-57.
9. Chang CC, Chow CC, Tellier LC, Vattikuti S, Purcell SM, Lee JJ. Second-generation PLINK: rising to the challenge of larger and richer datasets. Gigascience. 2015;4:7.
10. DerSimonian R, Laird N. Meta-analysis in clinical trials revisited. Contemp Clin Trials. 2015;45(Pt A):139-45.
11. McLaughlin T, Abbasi F, Cheal K, Chu J, Lamendola C, Reaven G. Use of metabolic markers to identify overweight individuals who are insulin resistant. Ann Intern Med. 2003;139(10):802-9.
12. Carpenter J, Bithell J. Bootstrap confidence intervals: when, which, what? A practical guide for medical statisticians. Stat Med. 2000;19(9):1141-64.
